# Relation of severe COVID-19 in Scotland to transmission-related factors and risk conditions eligible for shielding support: REACT-SCOT case-control study

**DOI:** 10.1101/2021.03.02.21252734

**Authors:** Paul M McKeigue, David A McAllister, David Caldwell, Ciara Gribben, Jen Bishop, Stuart McGurnaghan, Matthew Armstrong, Joke Delvaux, Sam Colville, Sharon Hutchinson, Chris Robertson, Nazir Lone, Jim McMenamin, David Goldberg, Helen M Colhoun

## Abstract

**Background:** Clinically vulnerable individuals have been advised to shield themselves during the COVID-19 epidemic. The objectives of this study were to investigate: (1) the risk of severe COVID-19 in those eligible for shielding, and (2) the relation of severe COVID-19 to transmission-related factors in those in shielding and the general population.

**Methods:** All 178578 diagnosed cases of COVID-19 in Scotland from 1 March 2020 to 18 February 2021 were matched for age, sex and primary care practice to 1744283 controls from the general population. This dataset (REACT-SCOT) was linked to the list of 212702 individuals identified as eligible for shielding. Severe COVID-19 was defined as cases that entered critical care or were fatal.

**Results:** With those without risk conditions as reference category, the univariate rate ratio for severe COVID-19 was 3.21 (95% CI 3.01 to 3.41) in those with moderate risk conditions and 6.3 (95% CI 5.8 to 6.8) in those eligible for shielding. The highest rate was in solid organ transplant recipients: rate ratio 13.4 (95% CI 9.6 to 18.8). Risk of severe COVID-19 increased with the number of adults but decreased with the number of school-age children in the household. Severe COVID-19 was strongly associated with recent exposure to hospital (defined as 5 to 14 days before presentation date): rate ratio 12.3 (95% CI 11.5 to 13.2) overall. To test for causality, a case-crossover analysis was undertaken; with less recent exposure only (15 to 24 days before first testing positive) as reference category, the rate ratio associated with recent exposure only was 5.9 (95% CI 3.6 to 9.7). The population attributable risk fraction for recent exposure to hospital peaked at 50% in May 2020 and again at 65% in December 2020.

**Conclusions:** The effectiveness of shielding vulnerable individuals was limited by the inability to control transmission in hospital and from other adults in the household. For solid organ transplant recipients, in whom the efficacy of vaccines is uncertain, these results support a policy of offering vaccination to household contacts. Mitigating the impact of the epidemic requires control of nosocomial transmission.

## Background

The SARS-CoV-2 pandemic reached Scotland in early 2020 with the first positive laboratory test recorded on 1 March 2020. Population-wide interventions included advice that symptomatic individuals should self-isolate, banning of mass gatherings, closure of schools and finally a population-wide lockdown on 23 March 2020. Although no systematic studies of risk factors were available at that time, public health agencies issued lists of “moderate risk” conditions [1] and “diseases and conditions considered to be very high risk” [2]. Those with “very high risk” conditions were designated as eligible for shielding, and were sent letters advising strict isolation and offering support which included a national opt-in scheme of free food delivery, home delivery of medication and priority access to supermarket delivery slots.

The initial objective of this study was to quantify the incidence of severe COVID-19 in the shielding population and to investigate whether shielding advice and support had reduced the risk of COVID-19. On the hypothesis that shielding was effective, we predicted that the rate ratio associated with eligibility for shielding compared to those without risk conditions would fall after the receipt of shielding letters. Our second objective was to understand the role of transmission-related factors including household composition and recent exposure to hospital on COVID-19 among those eligible for shielding. As the relevance of these transmission-related factors among those eligible for shielding became clear, the objectives were broadened to investigate the relation of severe COVID-19 to these transmission-related factors in the general population.

## Methods

We linked a national dataset of those eligible for shielding to a case-control dataset (REACT-SCOT) established in Public Health Scotland (PHS) that includes all COVID-19 cases in Scotland and matched population controls.[3] This case-control dataset is refreshed every few weeks and linked to health records that are used to assign a list of designated “moderate risk conditions”. Thus all cases and controls were classified into three categories: no risk condition, moderate risk condition only, eligible for shielding. The rate ratio of COVID-19 associated with each risk category was estimated as the conditional odds ratio as described below.

### Ascertainment of eligibility for shielding

The list of those eligible for shielding was generated by Public Health Scotland from March 2020 onwards by querying health-care information systems including hospital discharge records, prescription encashments, regional cancer chemotherapy databases, blood and transplant registries, for a designated list of diseases and conditions, supplemented direct requests to clinicians in relevant specialties [4]. The categories designated as eligible for shielding were as listed below [2,4]:

1. Solid organ transplant recipient
2. Cancer of the blood or bone marrow at any stage of treatment, or people with cancer receiving treatments that affect the immune system
3. Severe respiratory conditions including cystic fibrosis, severe asthma and severe chronic obstructive airway disease, on home oxygen, severe bronchiectasis, pulmonary hypertension)
4. Rare diseases that increase the risk of infections such as severe combined immunodeficiency, homozygous sickle-cell disease.
5. People on immunosuppression therapies sufficient to increase risk of infection
6. Pregnant with heart disease
7. Additional conditions, including people on renal dialysis, those who had had a splenectomy and others identified by clinicians as requiring shielding advice.

The first batch of shielding letters was sent on 3 April 2020. Further batches were issued on a weekly basis and the programme was paused on 1 August 2020. On 25 November 2020 a further letter was issued with “extra protection level advice for people at highest risk” based on the current protection level for the population level in that area. The list of those eligible for shielding has been regularly updated: this study is based on the list of 212702 individuals identified up to 28 January 2021.

### Ascertainment of cases and sampling of controls

Case ascertainment for the REACT-SCOT study has been described in detail elsewhere [3]. Case ascertainment was based on querying the following national-level databases: Electronic Communication of Surveillance in Scotland (ECOSS) that captures virology testing in all NHS laboratories, National Records of Scotland (NRS) death registrations, RAPID which is a daily update of hospitalisations, and Scottish Morbidity Record 01 (SMR01) which records general hospital discharges including day cases and is ICD-10 coded. All these databases use the Community Health Index (CHI) number as identifier. The CHI database includes age, sex, postcode and care home status, and can be queried to extract numbers of adults and children in the household.

Cases of COVID-19 were defined as those with a positive nucleic acid test for SARS-CoV-2 in ECOSS, a hospital discharge diagnosis of COVID-19 in SMR01, or a death registration with mention of COVID-19 anywhere on the death certificate. The presentation date was assigned as the date of the first positive test for those ascertained through testing, as seven days before the admission date for those without a positive test result ascertained through hospital discharge records, and as fourteen days before the date of death for those without a positive test result ascertained through death certificates. Databases were queried from 1 March 2020 (date of the first diagnosed case of COVID-19 in Scotland) up to 18 February 2021 for test results and 12 February 2021 for deaths.

Entry to critical care units – intensive care, high dependency or combined units – was obtained by linkage to the Scottish Intensive Care Society and Audit Group (SICSAG) database. Fatal outcome was defined as death at any time with COVID-19 coded as underlying cause, death from any cause within 28 days of testing positive or death within 28 days of presentation date for cases ascertained through discharge records. Severe COVID-19 was defined as entry to critical care within 21 days of presentation date, or fatal outcome. This definition was chosen to ensure that case ascertainment would not be affected by triage of those assessed as unlikely to benefit from critical care, or by the scale-up of SARS-CoV-2 testing.

For each case of COVID-19, up to 10 community controls matched for sex, one-year age band and primary care practice were selected from the CHI database, and assigned the same presentation date as the case. With this incidence density sampling design, it is possible and correct for an individual to appear more than once as a control and subsequently as a case.

### Linkage of cases and controls to demographic and morbidity data

Linkage of cases and controls to demographic and morbidity data and the associations of these factors with severe COVID-19 have been described in detail elsewhere [3]. Cases and controls were linked to hospital discharge ICD-10 codes over the last five years in SMR01, to British National Formulary codes of dispensed prescriptions in the 240 days before presentation date in the Prescribing Information System (PIS) and to the national register of diabetes. We used these linked datasets to assign a list of “moderate risk conditions” for COVID-19 designated by public health agencies [1]: diabetes, heart disease, asthma or chronic airway disease, chronic kidney disease, disabling neurological conditions, and immune deficiency or suppression. The codes used are as described previously [3]. Three broad risk groups were defined: no risk condition, moderate risk condition but ineligible for shielding, and eligible for shielding. Socioeconomic status was assigned as the Scottish Index of Multiple Deprivation (SIMD) score which is based on linkage of postcodes to Census data [5]; quintile 5 is the least deprived.

### Transmission-related risk factors

As addresses in the CHI database have been mapped to Unique Property Reference Numbers it was possible to calculate the numbers of adults and children in each household and to augment the coding of care home residence in the CHI database. Care home residence was assigned using the field in the CHI database, augmented by coding as care home residents the 4548 individuals aged over 70 in households with 10 or more adults to give a total of 26057 care home residents out of the 1922861 cases and controls. Linkage to occupational status for health-care workers and teachers was undertaken as described elsewhere [6,7].

We used the Scottish Morbidity Records SMR01 (inpatients and day cases) and SMR00 (outpatient attendance) together with the RAPID database to derive variables encoding recent exposure to hospitals. We defined the variable “recent hospital exposure” as any hospital in-patient stay, day case attendance or face-to-face out-patient consultation from 5 to 14 days before presentation date. Restriction of hospital exposure to this time window was intended to exclude consultations caused by COVID-19 symptoms for which testing was delayed by a few days, but to include those exposed to health care facilities in the time interval during which the infection was likely to have been acquired. Our intention was to capture all relevant exposure in cases and controls, rather than to assign cases as “health-care associated COVID-19” as specified by the European Centre for Disease Prevention and Control (ECDPC) [8]. The ECDPC case source definitions are restricted to cases who are already hospital in-patients when they first test positive: all other cases are classified as community onset.

### Statistical methods

The analyses presented here focus on severe COVID-19, defined as the main outcome measure of of the REACT-SCOT study at the design stage. Ascertainment of this outcome does not depend upon the extent of testing for SARS-CoV-2, which varied markedly over the study period and is likely to differ between those eligible for shielding and those ineligible. For the case-control analyses, cases and controls were censored at date of first vaccination and care home residents were excluded, as shielding advice intended for individuals in private households would not have been relevant to care home residents.

Cases and controls were grouped into three categories: no risk condition, moderate risk condition but ineligible for shielding, and eligible for shielding. Rate ratios for severe COVID-19 associated with eligibility for shielding are calculated with “no risk condition” as reference category. All rate ratios were estimated by conditional logistic regression. As the community controls were drawn by incidence density sampling and matched for age, sex, and general practice, this controls for these variables and for calendar time.

To plot the time course of the rate ratios associated with eligibility for shielding and with hospital exposure, these rate ratios were estimated over 21-day sliding windows of calendar time. The population attributable risk fraction (PARF) of severe cases for hospital exposure was calculated in each 21-day time window from Miettinen’s formula as *p*_*c*_ (*r −* 1) */r*. where *p*_*c*_ is the frequency of exposure in cases and *r* is the rate ratio [9].

Sliding windows of size 3, 7, and 7 days were used to plot the number of severe cases, the frequency of recent hospital exposure, and the rate ratios associated with hospital exposure and household composition. In the plots of exposure frequency and rate ratio the data points from 1 June 2020 to 30 September 2020 are omitted as the numbers of cases and controls are too low for estimates of frequencies and rate ratios to be accurate.

To test for causality of the association of recent exposure to hospital with severe COVID-19, we estimated rate ratios associated with recent hospital exposure (5 to 14 days before first testing positive) with less recent exposure (15 to 24 days before first testing positive) as reference category. This analysis was restricted to test-positive cases and their matched controls, so that it does not rely on the imputed presentation dates that were assigned to test-negative cases. The classic case-crossover design compares in cases only the frequencies of exposure in recent and less recent time windows, and estimates the conditional odds ratio as the ratio of number of cases with recent exposure only to number of cases with less recent exposure only. This can be viewed as a matched-pairs case-control study in which the case and control are the same person in recent and less recent time windows [10]. The analysis that we report is a refinement that takes advantage of the availability of a matched control group to contrast the conditional odds of being a case between those with recent exposure only and those with less recent exposure only. This controls for any difference in the population frequencies of exposure or disease incidence between recent and less recent time windows.

## Results

### Shielding eligibility and risk of COVID-19

Supplementary Table S1 shows the frequency of risk categories in those eligible for shielding. To comply with statistical disclosure control rules, the 58 women in the category “pregnant with heart disease” have been omitted from this table and all subsequent analyses; there were no severe cases in this group. Table 1 shows the number in each category of eligibility for shielding by case status classified as not diagnosed, not severe or severe COVID-19; among 212702 persons eligible for shielding 6081 (2.9%) were diagnosed with COVID-19 but did not enter critical care or die within 28 days, 359 (0.2%) entered critical care for COVID-19 but survived, and 1559 (0.7%) had a fatal outcome.

**Table 1.** Shielding eligibility cohort by case status. Severe cases are those that were fatal or entered critical care

|  | Not diagnosed<br>as case | No critical<br>care, non-fatal | Critical care,<br>non-fatal | Fatal | All |
| --- | --- | --- | --- | --- | --- |
| All shielding categories | 204703 (96.2%) | 6081 (2.9%) | 359 (0.2%) | 1559 (0.7%) | 212644 |
| **Shielding eligibility category** |  |  |  |  |  |
| Solid organ transplant | 6620 (96.3%) | 188 (2.7%) | 24 (0.3%) | 40 (0.6%) | 6872 |
| Specific cancers | 25788 (96.6%) | 652 (2.4%) | 36 (0.1%) | 223 (0.8%) | 26699 |
| Severe respiratory | 83268 (96.2%) | 2432 (2.8%) | 131 (0.2%) | 768 (0.9%) | 86599 |
| Rare diseases | 10673 (96.4%) | 300 (2.7%) | 14 (0.1%) | 85 (0.8%) | 11072 |
| On immunosuppressants | 30796 (96.8%) | 866 (2.7%) | 42 (0.1%) | 103 (0.3%) | 31807 |
| Additional conditions | 47501 (95.8%) | 1642 (3.3%) | 112 (0.2%) | 340 (0.7%) | 49595 |

Percentages are row percentages

As shown in Figure 1 the time course of daily severe cases in those eligible for shielding paralleled the time course in those without risk conditions. However in the first wave the daily number of severe cases fell more slowly in those eligible for shielding than in those without risk conditions. Of the 1926 severe cases among those eligible for shielding, 286 were resident in care homes. In the case-control analyses, care home residents were excluded, as policies for shielding residents of care homes are different from those relevant to shielding individuals living independently.

**Fig 1.**
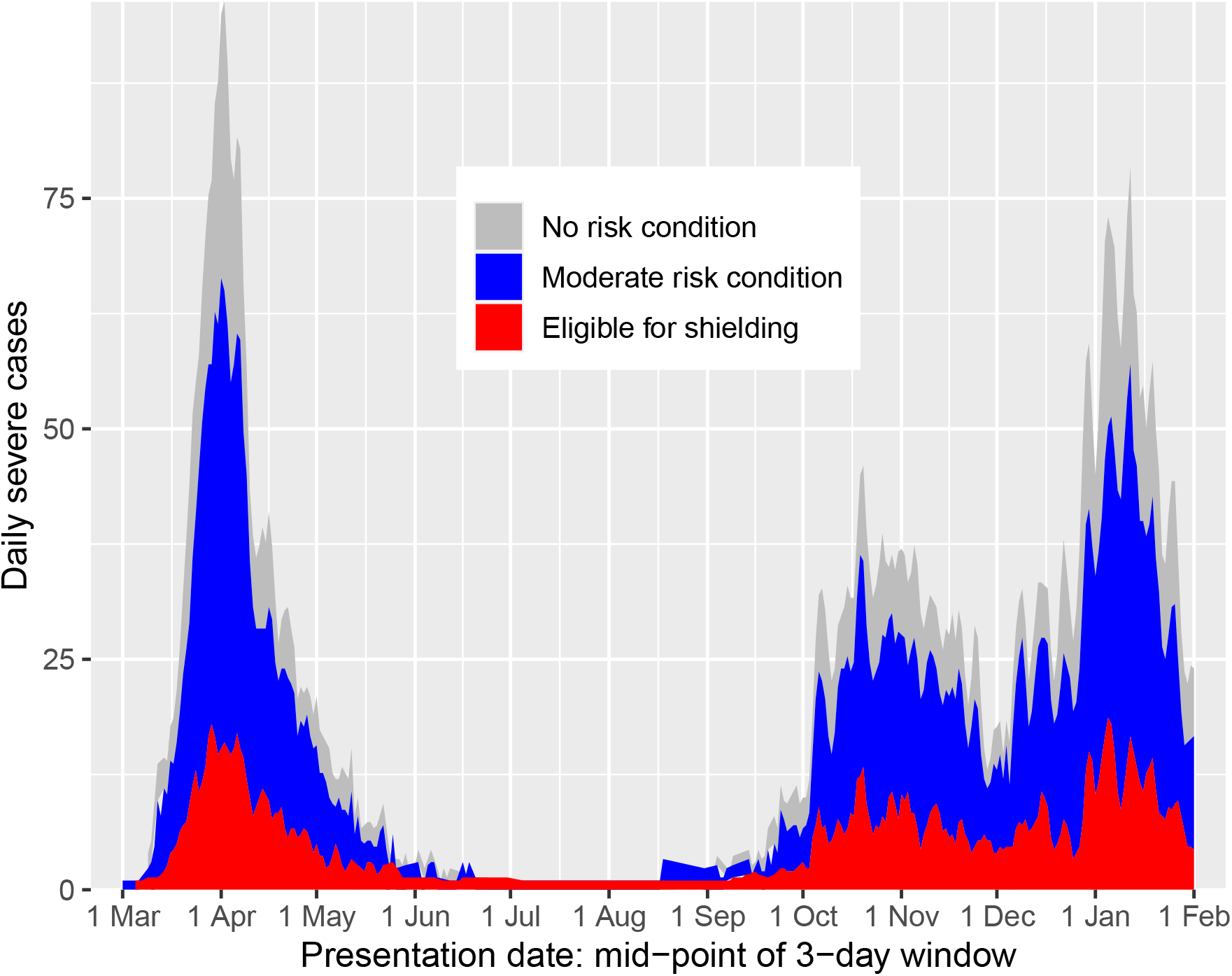
Area plot of severe cases by vulnerability category and date of presentation, excluding care home residents

### Rate ratios for severe COVID-19 by risk group

Table 2 shows the rate ratios for severe COVID-19 associated with each category of shielding eligibility, with those without risk conditions as reference category, excluding care home residents. The univariate rate ratio for severe disease was 3.21 (95% CI 3.01 to 3.41, *p*=2 *×* 10^*−*298^) in those with moderate risk conditions and 6.3 (95% CI 5.8 to 6.8, *p*=3 *×* 10^*−*479^) in those eligible for shielding. Among those eligible for shielding, solid organ transplant recipients were the group at highest risk, with a univariate rate ratio of 13.4 (95% CI 9.6 to 18.8, *p*=2 *×* 10^*−*51^) for severe COVID-19. On multivariate adjustment rate ratios for all these groups remained high.

**Table 2.** Rate ratios for severe COVID-19 in those not resident in care homes.

|  | Controls<br>(86902) | Cases (7217) | Univariate |  | Multivariable |  |
| --- | --- | --- | --- | --- | --- | --- |
|  |  |  | Rate ratio (95%<br>CI) | *p*-value | Rate ratio (95%<br>CI) | *p*-value |
| No risk condition | 45354 (52%) | 1897 (26%) |  | . |  | . |
| Moderate risk condition | 33909 (39%) | 3702 (51%) | 3.20 (3.01, 3.41) | $2 \times 10^{-297}$ | 2.67 (2.50, 2.85) | $2 \times 10^{-184}$ |
| Shielding eligibility category |  |  |  |  |  |  |
| Solid organ transplant | 112 (0%) | 59 (1%) | 13.4 (9.6, 18.8) | $2 \times 10^{-51}$ | 7.0 (4.6, 10.4) | $5 \times 10^{-21}$ |
| Specific cancers | 907 (1%) | 240 (3%) | 7.9 (6.8, 9.3) | $8 \times 10^{-143}$ | 3.47 (2.87, 4.19) | $1 \times 10^{-37}$ |
| Severe respiratory | 3751 (4%) | 733 (10%) | 5.8 (5.2, 6.4) | $2 \times 10^{-263}$ | 4.32 (3.87, 4.83) | $3 \times 10^{-149}$ |
| Rare diseases | 341 (0%) | 69 (1%) | 7.0 (5.3, 9.3) | $1 \times 10^{-40}$ | 4.68 (3.37, 6.49) | $2 \times 10^{-20}$ |
| On immunosuppressants | 742 (1%) | 126 (2%) | 4.61 (3.76, 5.65) | $3 \times 10^{-49}$ | 3.26 (2.59, 4.10) | $8 \times 10^{-24}$ |
| Additional conditions | 1786 (2%) | 391 (5%) | 6.6 (5.8, 7.5) | $3 \times 10^{-180}$ | 4.31 (3.72, 5.00) | $3 \times 10^{-84}$ |
| At least one child under 5 | 1759 (2%) | 150 (2%) | 0.87 (0.73, 1.04) | 0.1 | 0.73 (0.60, 0.89) | 0.002 |
| Number of school age children in household |  |  |  |  |  |  |
| 0 school age | 80885 (93%) | 6685 (93%) |  | . |  | . |
| 1 school age | 1749 (2%) | 93 (1%) | 0.54 (0.43, 0.67) | $4 \times 10^{-8}$ | 0.76 (0.60, 0.96) | 0.02 |
| 2 or more | 4268 (5%) | 439 (6%) | 1.06 (0.95, 1.19) | 0.3 | 0.71 (0.62, 0.80) | $2 \times 10^{-7}$ |
| Number of adults in household |  |  |  |  |  |  |
| 1 adult | 62011 (72%) | 4259 (59%) |  | . |  | . |
| 2 adults | 19190 (22%) | 1951 (27%) | 1.57 (1.48, 1.67) | $1 \times 10^{-49}$ | 1.80 (1.68, 1.92) | $8 \times 10^{-65}$ |
| 3 to 4 | 4611 (5%) | 769 (11%) | 2.50 (2.28, 2.74) | $3 \times 10^{-88}$ | 3.19 (2.87, 3.54) | $6 \times 10^{-104}$ |
| 5 to 9 | 326 (0%) | 85 (1%) | 3.98 (3.10, 5.11) | $3 \times 10^{-27}$ | 5.8 (4.3, 7.7) | $1 \times 10^{-33}$ |
| 10 or more | 247 (0%) | 102 (1%) | 10.6 (7.6, 14.6) | $1 \times 10^{-45}$ | 11.4 (8.0, 16.2) | $1 \times 10^{-40}$ |
| SIMD quintile |  |  |  |  |  |  |
| 1 - most deprived | 21388 (25%) | 2162 (30%) |  | . |  | . |
| 2 | 19072 (22%) | 1761 (24%) | 0.91 (0.85, 0.98) | 0.01 | 0.95 (0.88, 1.03) | 0.3 |
| 3 | 15399 (18%) | 1264 (18%) | 0.76 (0.70, 0.82) | $5 \times 10^{-11}$ | 0.84 (0.76, 0.92) | $1 \times 10^{-4}$ |
| 4 | 14870 (17%) | 1079 (15%) | 0.65 (0.59, 0.70) | $1 \times 10^{-22}$ | 0.72 (0.65, 0.79) | $3 \times 10^{-11}$ |
| 5 - least deprived | 16084 (19%) | 944 (13%) | 0.53 (0.48, 0.58) | $3 \times 10^{-38}$ | 0.60 (0.53, 0.66) | $1 \times 10^{-20}$ |
| Occupation |  |  |  |  |  |  |
| Other / undetermined | 84672 (98%) | 6962 (97%) |  | . |  | . |
| Teacher | 336 (0%) | 11 (0%) | 0.35 (0.19, 0.64) | $7 \times 10^{-4}$ | 0.42 (0.22, 0.81) | 0.01 |
| Health care, not PF / undetermined | 826 (1%) | 87 (1%) | 1.18 (0.94, 1.47) | 0.2 | 1.20 (0.94, 1.53) | 0.2 |
| Health care PF | 846 (1%) | 145 (2%) | 1.80 (1.50, 2.16) | $3 \times 10^{-10}$ | 1.86 (1.52, 2.27) | $2 \times 10^{-9}$ |
| Recent hospital visit/stay | 3565 (4%) | 2415 (33%) | 12.3 (11.5, 13.2) | $3 \times 10^{-1106}$ | 10.2 (9.5, 11.0) | $2 \times 10^{-824}$ |

Percentages are column percentages for each variable
PF, patient-facing
Rate ratios are from conditional logistic regression models with cases and controls matched for age, sex and general care practice
Univariate rate ratios are for models with a single covariate
Multivariable rate ratios are for a model including all covariates

We examined the time course of the rate ratio associated with eligibility for shielding. As shown in Supplementary Table S3 most of the solid organ transplant recipients, those with severe respiratory disease, and cancer patients were included in the first batch of shielding letters sent on 3 April 2020. Supplementary Figure S1(a) shows the risk ratio associated with eligibility for shielding increased from 5.17 in the time window with mid-point 1 April to 8.89 in the time window with mid-point 1 May. The rate ratio associated with moderate risk conditions also rose in the first half of May, but fell more rapidly than the rate ratio associated with eligibility for shielding. Thus there was no evidence that shielding advice reduced the rate ratio.

## Associations of severe COVID-19 with transmission-related factors

### Associations in those eligible for shielding

Table 3 shows associations with risk factors among those eligible for shielding only, with severe respiratory disease (the largest category) as reference category. The rate ratio increased with the number of adults in the household but not with the number of children. The strongest risk factor for severe COVID-19 among those eligible for shielding was recent exposure to hospital, with a rate ratio of 6.0 (95% CI 4.7 to 7.7, *p*=1 *×* 10^*−*46^) in the multivariable model. Of severe cases among those eligible for shielding, 739 (45%) had recent exposure to hospital. Using Miettinen’s formula as given above, the PARF for recent exposure to hospital in those eligible for shielding can thus be calculated as 37% of severe cases.

**Table 3.**
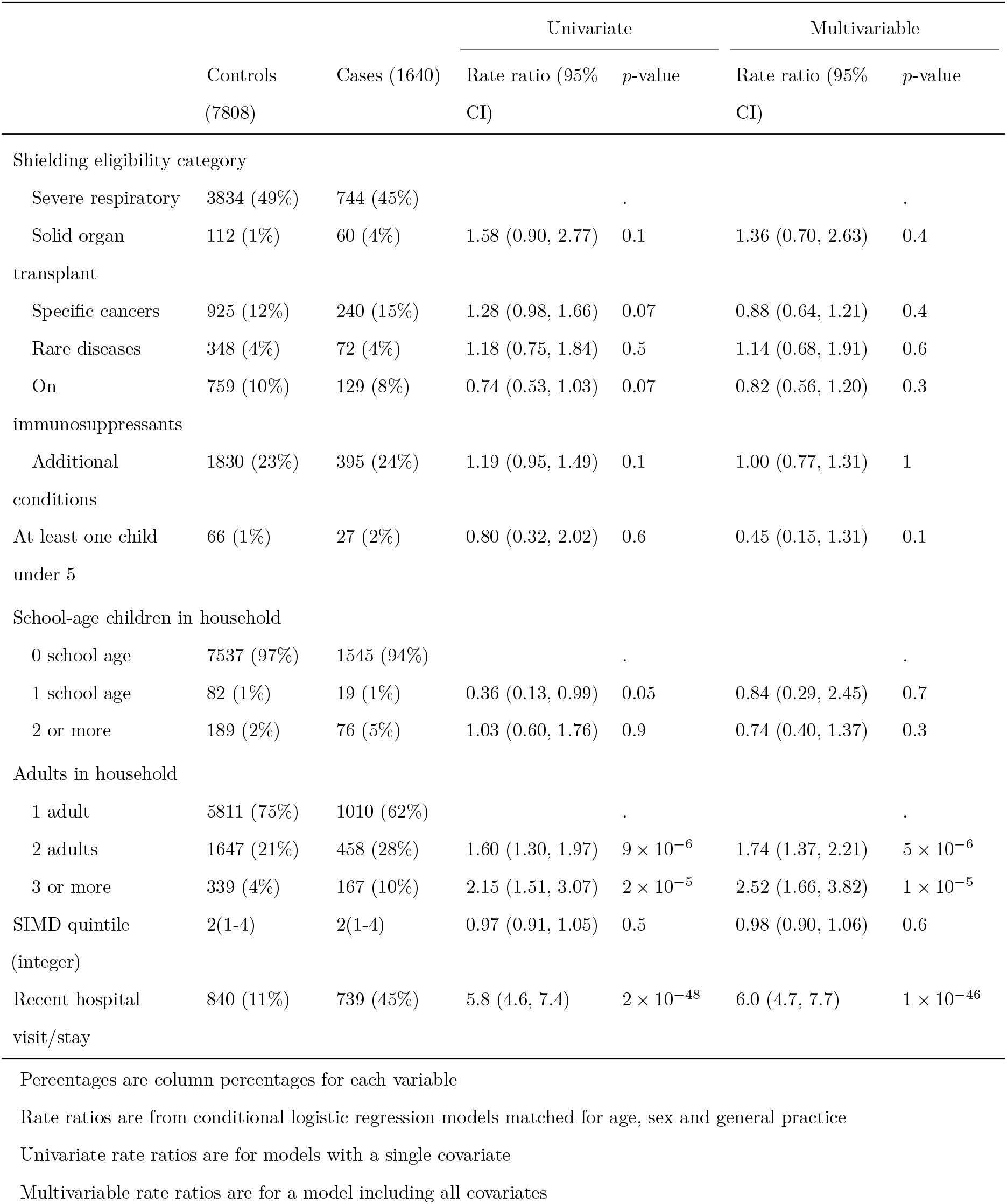
Rate ratios for severe COVID-19 associated with risk conditions (with severe respiratory disease as reference category) in those eligible for shielding and not resident in care homes

|  |  |  | Univariate |  | Multivariable |  |
| --- | --- | --- | --- | --- | --- | --- |
|  | Controls | Cases (1640) | Rate ratio (95% | *p*-value | Rate ratio (95% | *p*-value |
|  | (7808) |  | CI) |  | CI) |  |
| Shielding eligibility category |  |  |  |  |  |  |
| Severe respiratory | 3834 (49%) | 744 (45%) |  | . |  | . |
| Solid organ transplant | 112 (1%) | 60 (4%) | 1.58 (0.90, 2.77) | 0.1 | 1.36 (0.70, 2.63) | 0.4 |
| Specific cancers | 925 (12%) | 240 (15%) | 1.28 (0.98, 1.66) | 0.07 | 0.88 (0.64, 1.21) | 0.4 |
| Rare diseases | 348 (4%) | 72 (4%) | 1.18 (0.75, 1.84) | 0.5 | 1.14 (0.68, 1.91) | 0.6 |
| On immunosuppressants | 759 (10%) | 129 (8%) | 0.74 (0.53, 1.03) | 0.07 | 0.82 (0.56, 1.20) | 0.3 |
| Additional conditions | 1830 (23%) | 395 (24%) | 1.19 (0.95, 1.49) | 0.1 | 1.00 (0.77, 1.31) | 1 |
| At least one child under 5 | 66 (1%) | 27 (2%) | 0.80 (0.32, 2.02) | 0.6 | 0.45 (0.15, 1.31) | 0.1 |
| School-age children in household |  |  |  |  |  |  |
| 0 school age | 7537 (97%) | 1545 (94%) |  | . |  | . |
| 1 school age | 82 (1%) | 19 (1%) | 0.36 (0.13, 0.99) | 0.05 | 0.84 (0.29, 2.45) | 0.7 |
| 2 or more | 189 (2%) | 76 (5%) | 1.03 (0.60, 1.76) | 0.9 | 0.74 (0.40, 1.37) | 0.3 |
| Adults in household |  |  |  |  |  |  |
| 1 adult | 5811 (75%) | 1010 (62%) |  | . |  | . |
| 2 adults | 1647 (21%) | 458 (28%) | 1.60 (1.30, 1.97) | $9 \times 10^{-6}$ | 1.74 (1.37, 2.21) | $5 \times 10^{-6}$ |
| 3 or more | 339 (4%) | 167 (10%) | 2.15 (1.51, 3.07) | $2 \times 10^{-5}$ | 2.52 (1.66, 3.82) | $1 \times 10^{-5}$ |
| SIMD quintile (integer) | 2(1-4) | 2(1-4) | 0.97 (0.91, 1.05) | 0.5 | 0.98 (0.90, 1.06) | 0.6 |
| Recent hospital visit/stay | 840 (11%) | 739 (45%) | 5.8 (4.6, 7.4) | $2 \times 10^{-48}$ | 6.0 (4.7, 7.7) | $1 \times 10^{-46}$ |

Percentages are column percentages for each variable
Rate ratios are from conditional logistic regression models matched for age, sex and general practice
Univariate rate ratios are for models with a single covariate
Multivariable rate ratios are for a model including all covariates

### Associations in the overall population

Table 2 shows the association of severe COVID-19 with risk factors in the general population, including those eligible for shielding but excluding residents in care homes. The risk of severe COVID-19 increased with the number of adults in the household, but was inversely associated with the number of school-age children in the household in a multivariable model. The rate ratio associated with two or more adults (with single-adult households as reference category), was 2.08 (95% CI 1.95 to 2.21, *p*=1 *×* 10^*−*116^) and the rate ratio associated with one or more school-age children was 0.72 (95% CI 0.64 to 0.81, *p*=3 *×* 10^*−*8^). The other demographic factor associated with increased risk was socioeconomic deprivation: the rate ratio in the least deprived quintile compared with the most deprived quintile was 0.60 (95% CI 0.53 to 0.66, *p*=1 *×* 10^*−*20^). As shown in Supplementary Figure S1(b), in a joint model with number of adults in household, number of children in household, and SIMD deprivation score the rate ratio for severe disease per adult in household remained in the range 1.5 to 2 throughout the epidemic, and the rate ratio per school-age child in household remained mostly in the range 0.7 to 1.

As reported previously, in comparison with other occupations patient-facing health-care workers were at higher risk of severe disease with a rate ratio of 1.80 (95% CI 1.50 to 2.16, *p*=3 *×* 10^*−*10^), and teachers were at lower risk with a rate ratio of 0.35 (95% CI 0.19 to 0.64, *p*=7 *×* 10^*−*4^).

### Association with recent exposure to hospital

As Table 2 shows, the strongest risk factor (in terms of deviance explained) for severe COVID-19 in the overall population was recent exposure to hospital: rate ratio 12.3 (95% CI 11.5 to 13.2, *p*=3 *×* 10^*−*1106^). Exclusion of the 11% of those dying within 28 days of a positive test who did not have COVID-19 as underlying cause of death on their death certificates changed this rate ratio only slightly to 11.9 (11.0, 12.8).

Figure 2(a) shows the time course of the frequency of recent exposure to hospital using data from controls; for monitoring background exposure levels it is appropriate to examine the frequency in controls rather than in cases. Recent exposure to hospital fell precipitously when restrictions on non-COVID-19 admission were imposed in March, but remained higher in those eligible for shielding than in those with moderate risk conditions or no risk conditions throughout.

**Fig 2.**
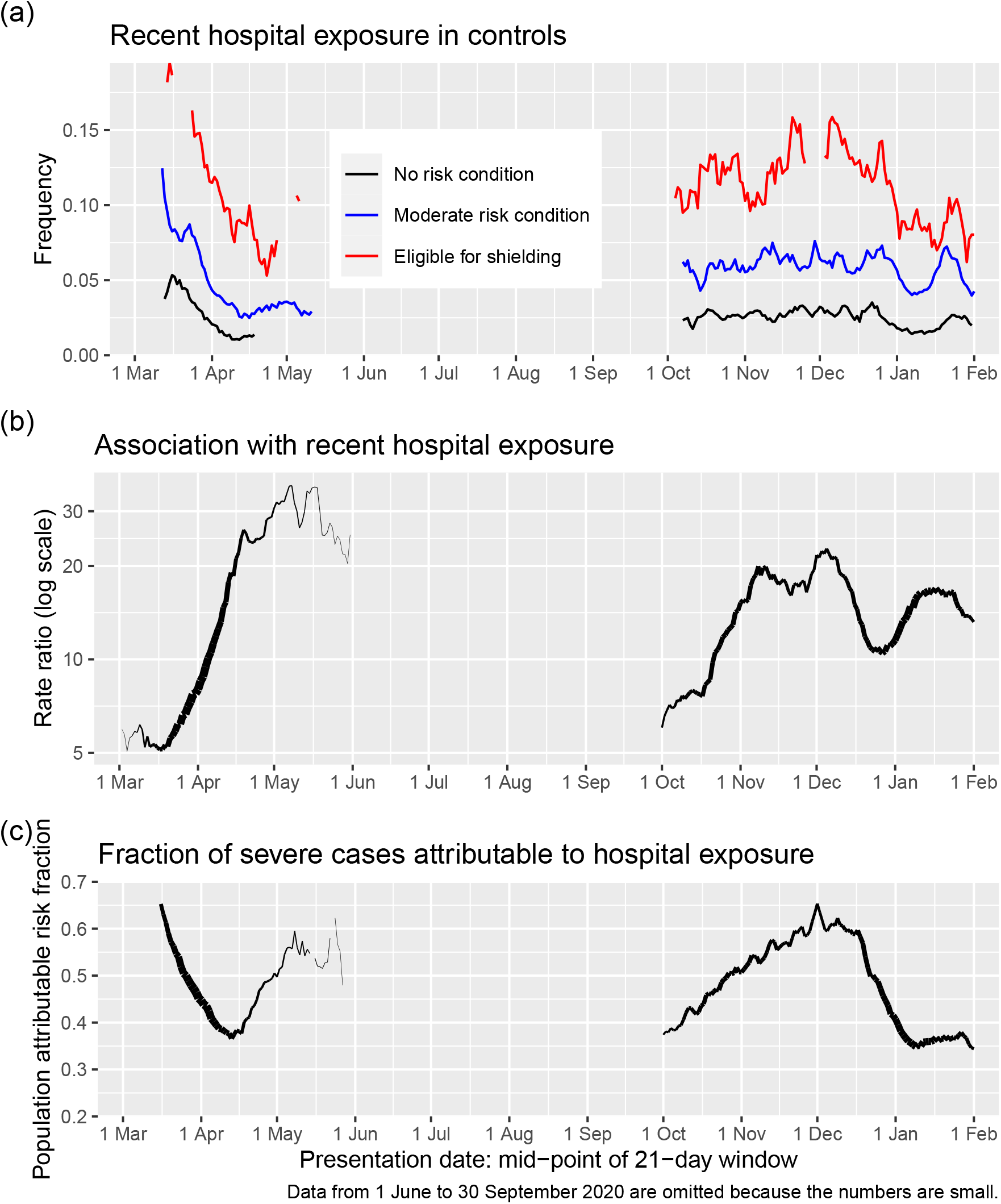
(a) Recent hospital exposure by risk group and sliding window of presentation dates (b) Rate ratio asociated with recent hospital exposure in those not resident in care homes. (c) fraction of severe cases attributable to recent hospital exposure in those not resident in care homes.

As shown in Figure 2(b) the rate ratio associated with recent exposure to hospital increased from 8 in the time window with mid-point 1 April to a rate ratio of 31 in the time window with mid-point 1 May.

The PARF of severe COVID-19 for recent exposure to hospital in the general population can be calculated as 30% over the study period. This fraction reached a peak in the first wave of 50% of severe cases at the beginning of May, declined rapidly over the next few weeks, and increased again to 65% of severe cases at the beginning of December (Figure 2(c)).

In a *post hoc* analysis we found that the association of recent exposure to hospital with severe disease was largely driven by inpatient exposure [rate ratio 33.6 (95% CI 30.4 to 37.3, *p*=7 *×* 10^*−*984^)]; the rate ratios associated with day case exposure [2.59 (95% CI 1.87 to 3.58, *p*=9 *×* 10^*−*9^)] or outpatient exposure [2.67 (95% CI 2.40 to 2.97, *p*=4 *×* 10^*−*71^)] were much lower. To test if the association of severe COVID-19 with recent inpatient exposure was likely to simply reflect confounding by time invariant comorbidity, the association was examined by time window as described in the Methods section. Table 4 shows that the rate ratio for severe COVID-19 associated with inpatient exposure only in the recent interval (days 5 to 14 before testing positive), with exposure only in the less recent interval (days 15 to 24 before testing positive) as reference category was 5.9 (95% CI 3.6 to 9.7, *p*=3 *×* 10^*−*12^).

**Table 4.** Rate ratios for severe COVID-19 by time window of hospital in-patient exposure

| Time window of exposure | Controls | Cases | Rate ratio (95%<br>CI) | *p*-value |
| --- | --- | --- | --- | --- |
| Less recent interval only | 494 (29%) | 147 (6%) |  | . |
| Recent interval only | 470 (28%) | 799 (35%) | 5.9 (3.6, 9.7) | $3 \times 10^{-12}$ |
| Both intervals | 741 (43%) | 1335 (59%) | 6.9 (4.3, 11.1) | $3 \times 10^{-15}$ |

Rate ratios are from conditional logistic regression models
Dataset restricted to those exposed at some time between 5 and 24 days before first testing positive.
Recent interval = days 5 to 14, less recent interval days 15 to 24 before first testing positive.
Reference category is exposure in less recent interval only.

The frequency of recent inpatient exposure over the period of study in cases not resident in care homes was 28%. From this and the rate ratio of 33.6 given above the PARF for recent hospital inpatient exposure can be calculated from Miettinen’s formula as 27%. As Table S2, shows, most of the difference in inpatient exposure classification of severe cases between our definition and the ECDPC definition arises because cases with onset after hospital discharge would be classified as community onset by the ECDPC classification: only 57% of severe cases with inpatient exposure 5-14 days before COVID-19 presentation would be defined as probable or definite hospital-acquired infection (in hospital for at least 8 days before first testing positive) by the ECPDC classification. The rate ratio for severe disease associated with probable or definite hospital-acquired infection as defined by the ECDPC classification was 45.4.

## Discussion

### Summary of findings and comparison with other studies

Key findings of this study were: (1) that the rate ratios for severe COVID-19 associated with shielding conditions remained high through the epidemic; (2) that recent hospital exposure and the number of adults in the household were associated with severe COVID-19 among those eligible for shielding and also in the general population; (3) that among those not resident in care homes the proportion of severe cases attributable to hospital attendance reached a peak of 65% during the second wave.

### Risk to those eligible for shielding

In comparison with those with no risk condition, the highest rate ratio for severe COVID-19 was the 13-fold rate ratio in solid organ transplant recipients. For other conditions deemed eligible for shielding, the rate ratios were between 5 and 8, compared to the rate ratio of about 3 associated with conditions designated as moderate risk, which include heart disease, diabetes, chronic kidney disease and disabling neurological conditions. The numbers of pregnant women with heart disease were too small for the risk in this group to be estimated. Other studies of outcome of COVID-19 in solid organ transplant recipients and other individuals using immunosuppressants have been based only on patients admitted to hospital [11,12]. Such studies cannot assess the risks to immunosuppressed individuals in the population.

If the advice and support offered in the shielding letters had been effective (beyond the messages already widely disseminated by this time and general social distancing measures that were introduced by then), we would have expected the rate ratio for severe disease associated with eligibility for shielding to fall within two weeks of the first letters being sent out. Our results show however that while presentations of severe cases fell rapidly in the general population from the beginning of April 2020, the fall in presentations among those eligible for shielding advice was delayed so that the rate ratio for severe COVID-19 associated with eligibility for shielding rose during April 2020, although the daily deaths fell during April 2020 both in those eligible and those ineligible for shielding. The rate ratios associated with eligibility for shielding and with moderate risk conditions rose again from October 2020 to early December.

The high frequency of recent hospital exposure in cases of severe COVID-19 who were eligible for shielding, together with the increase in the rate ratio associated with recent hospital exposure during periods when population-wide social distancing measures were being imposed, suggests that exposure to transmission in hospital settings is at least part of the explanation for the failure to reduce the rate ratio associated with eligibility for shielding. Our finding that even in those advised to shield the risk of severe disease was increased in those who were sharing a household with other adults suggests that this was another constraint on the effectiveness of advice to shield, as no support was provided for other household members to co-isolate with the vulnerable individual.

### Association with household composition

The increased risk of severe COVID-19 in patient facing health care workers is consistent with our previous report of a threefold risk for hospitalised COVID-19 earlier in the epidemic [6]. We and others have previously reported lower risk in teachers compared with others of the same age and sex [7,13]. The inverse association of severe disease with the number of school-age children in the household extends and confirms the findings of an earlier study of health care workers and their families [14]. In the OPENSAFELY cohort, the rate ratio for fatal COVID-19 associated with living with children aged 0-11 years was 0.75 after adjusting for covariates, but no dose-response relationship was reported [15]. The inverse association of severe COVID-19 with past exposure to children is consistent with evidence that other coronaviruses generate cross-reactive T-cell responses that may confer some resistance to SARS-CoV-2 [16]. From a public health perspective the most relevant implication is that although the rate ratio per child has been higher in the second wave of the epidemic than in the first wave, it has remained below 1 in almost all time windows. The association of severe COVID-19 with number of adults in the household is consistent with classic studies of other viral infections showing that secondary cases in a household, where the infecting dose is likely to be high, are more severe than index cases [17].

### Association with recent exposure to hospital

A striking finding from our analysis is the PARF of 30 % for severe disease associated with recent exposure to hospital. Although we had pre-specified this category to include day case and outpatient exposure, the association was driven by inpatient exposure. The PARF associated with inpatient exposure was 27%. Using the accepted international definitions of probable or definite nosocomial acquisition gives a much lower estimate of the PARF. As we have shown most of this difference is due to the fact that the international definition does not capture cases diagnosed shortly after discharge that could have been acquired in hospital. The other difference is that the international definition classifies those with onset 5-6 days after admission as indeterminate. As the average incubation period for COVID-19 is 5-6 days, exposure in this time window is relevant.

The role of nosocomial transmission was recognized early in the epidemic. Hospital-acquired infection was suspected in 41% of hospitalised cases seen in January 2020 in one centre in Wuhan [18]. A commentary in April 2020 noted that “in Lombardy, SARS-CoV-2 became largely a nosocomial infection” [19]. Most previous reports have focused on the narrow definition of probable hospital-acquired infection, rather than the association of disease with exposure to hospital [20]. The ARHAI (Antimicrobial Resistance and Healthcare Associated Infection) unit of NHS National Services Scotland recently reported that among those who died within 28 days of testing positive for COVID-19 (including care home residents), 1015 met the ECDPC definition of possible or probable hospital-acquired infection [21].

A paper presented to SAGE on 28 January 2021 broadened the ECDPC definition of health-care associated infection to include cases discharged from hospital up to 14 days before first testing positive as “community-onset suspected healthcare-associated”, similar to the definition that we have used for “recent hospital exposure” [22]. From linking hospital episode data to COVID-19 tests and CO-CIN, the authors estimated that 40% of cases hospitalised with COVID-19 were exposed by this broad definition, and that onward transmission of COVID-19 from hospital-acquired cases could account for another 5% of hospitalised cases.

### Strengths and limitations

Strengths of our study are the national coverage and the comprehensive linkage to medical records and demographic risk factors. A limitation is that we do not have primary care data other than encashed prescriptions. Furthermore, as most immunosuppressant drugs are prescribed through hospitals where linkage to prescribing records is not yet possible, the risks associated with specific immunosuppressant drug classes could not be investigated. We have no data on help with daily activities from non-resident carers as another possible source of exposure of clinically vulnerable individuals attempting to shield themselves.

The calculation of the PARF for an exposure provides an upper bound on the predicted effect of removing that exposure. The definition of “recent hospital exposure” used in this study was intended to capture all those whose infection could have been acquired in hospital (sensitivity of 1), unlike the international definition of “probable health-care associated infection” which is intended to identify those whose infection was unlikely to have been acquired outside hospital (high specificity). This inclusive definition is appropriate, as the calculation of the PARF is valid if the classification of exposure has sensitivity of 1 even if the specificity is less than 1 [9]. The association of severe COVID-19 with recent hospital admission is likely to be confounded by pre-existing risk conditions. However adjusting for risk conditions in a multivariable analysis reduced the rate ratio associated with recent hospital exposure only slightly (from 12.3 to 10.2). The most compelling evidence of causality is the time window analysis, which shows that the association of disease is with elapsed time since exposure corresponding to the known incubation period of SARS-CoV-2. The rate ratio estimated from the time window analysis is not directly comparable with the rate ratio based on the case-control analysis because restriction to those with discordant exposure between time windows selects those with fewer days in hospital (lower dosage of the exposure).

As all hospital inpatients are tested for SARS-CoV-2 every few days and all deaths within 28 days of a positive test are officially classified as deaths involving COVID-19, some misclassification of deaths from other causes as COVID-19 deaths is likely in those with recent hospital exposure. However the rate ratio associated with recent hospital exposure was barely changed when fatal cases without COVID-19 as underlying cause of death on death certificate were omitted.

### Policy implications

Our results have implications for policies on shielding the vulnerable, vaccination, and control of SARS-CoV-2 transmission in the population.

We found no evidence that the shielding programme *per se* reduced COVID-19 rates although it is possible that without shielding advice and support the outcome in this group would have been worse. It is relevant to examine why advice to shield, combined with offers of support for delivery of food and medicines, failed to protect some individuals who were identified as clinically extremely vulnerable. As a lockdown on the general population had been imposed on 23 March 2020, and individuals who considered themselves to be at high risk would have been likely to reduce their contact level before then, there were limited possibilities for risk in this group to be reduced further by advice to shield and offers of help in letters sent from 3 April onwards. We have identified two sources of exposure that are associated with severe disease and cannot easily be avoided by those advised to shield: in-patient hospital care, and sharing a household with other adults. We recommend that consideration should be given to special measures for to allow these vulnerable individuals to reduce their exposure. This could include special measures to protect these individuals from nosocomial infection, and support for other household members to co-isolate with the vulnerable individual.

Our findings support the policy of assigning highest priority for vaccination to those with risk conditions eligible for shielding, as these groups have markedly elevated risks of severe COVID-19. For solid organ transplant recipients – the group at highest risk – it is uncertain whether the vaccine will evoke an immune response sufficient to be effective [23]. As there is now evidence that vaccination reduces transmission to household contacts [24], one way to reduce risk in this group would be to vaccinate their household contacts. Even with vaccination of household contacts, solid organ transplant recipients will require shielding until passive immunisation is available or the risk of transmission has been reduced to a very low level by herd immunity.

Policies for control of transmission in the population and for reducing burden on health services and total deaths have focused on population-wide reduction of social contact. However our analysis provides compelling evidence for a substantial contribution of nosocomial transmission to the burden of severe COVID-19 even during the second wave. A report for NHS England by the Healthcare Safety Investigation Branch noted the challenges of controlling nosocomial transmission and recommended that a national strategy for infection prevention and control should be developed [25]. Vaccination of health care workers is likely to reduce nosocomial transmission, but vaccination of those booked for elective procedures should also be considered. We have not attempted to quantify the onward transmission of COVID-19 from discharged patients, but the PHE modelling study estimated that this could have accounted for 5% of cases requiring hospitalisation in England during the first wave [22]. This suggests more stringent testing before discharge and quarantine post discharge should be considered. More detailed understanding of how recommended infection control policies are being operationalised is also needed [26].

## Data Availability

The component datasets used here are available via the Public Benefits Privacy Panel for Health at
https://www.informationgovernance.scot.nhs.uk/pbpphsc/ for researchers who meet the criteria for access to confidential data. All source code used for derivation of variables, statistical analysis and generation of this manuscript is available on https://github.com/pmckeigue/covid-scotland_public

## Declarations

### Ethics approval and information governance

This study was approved under COVID-19 Rapid Data Protection Impact Assessment (DPIA) 20210023 that allows Public Health Scotland staff to link the datasets. Datasets were de-identified before analysis.

### Data Availability

The component datasets used here are available via the Public Benefits Privacy Panel for Health at https://www.informationgovernance.scot.nhs.uk/pbpphsc/ for researchers who meet the criteria for access to confidential data. All source code used for derivation of variables, statistical analysis and generation of this manuscript is available on https://github.com/pmckeigue/covid-scotland_public.

### Registration

The original protocol for the REACT-SCOT case-control study was registered with the European Network of Centres for Pharmacoepidemiology and Pharmacovigilance (ENCEPP number EUPAS35558).

## Acknowledgements

We thank all staff in critical care units who submitted data to the SICSAG database, the Scottish Morbidity Record Data Team, the staff of the National Register of Scotland, the Public Health Scotland Terminology Services, the HPS COVID-19 Laboratory & Testing cell and the NHS Scotland Diagnostic Virology Laboratories, and Nicola Rowan (HPS) for coordinating this collaboration. We thank Shona Cairns and Laura Imrie of the Antimicrobial Resistance and Healthcare Associated Infection unit of NHS National Services Scotland for comments on an earlier draft of this manuscript.

## Public Health Scotland COVID-19 Health Protection Study Group

Allan McLeod, Alice Whettlock April Went, Beth Findlay Chris Sullivan, Ciaran Harvey, David Henderson, Edward McArdle, Eisin McDonald, Emily Griffiths, Genna Drennan, Johanna Young, Kirstin Leslie, Leonardo I Green, Louise Nicol, Melissa Llano, Nick Christofidis, Paul Bett, Ross Cameron, Theresa Ryan, Victoria Ponce Hardy.

Public Health Scotland, Meridian Court, 5 Cadogan Street, Glasgow G2 6QE

## Supplementary Material

### Supplementary Figures

**Fig S1.**
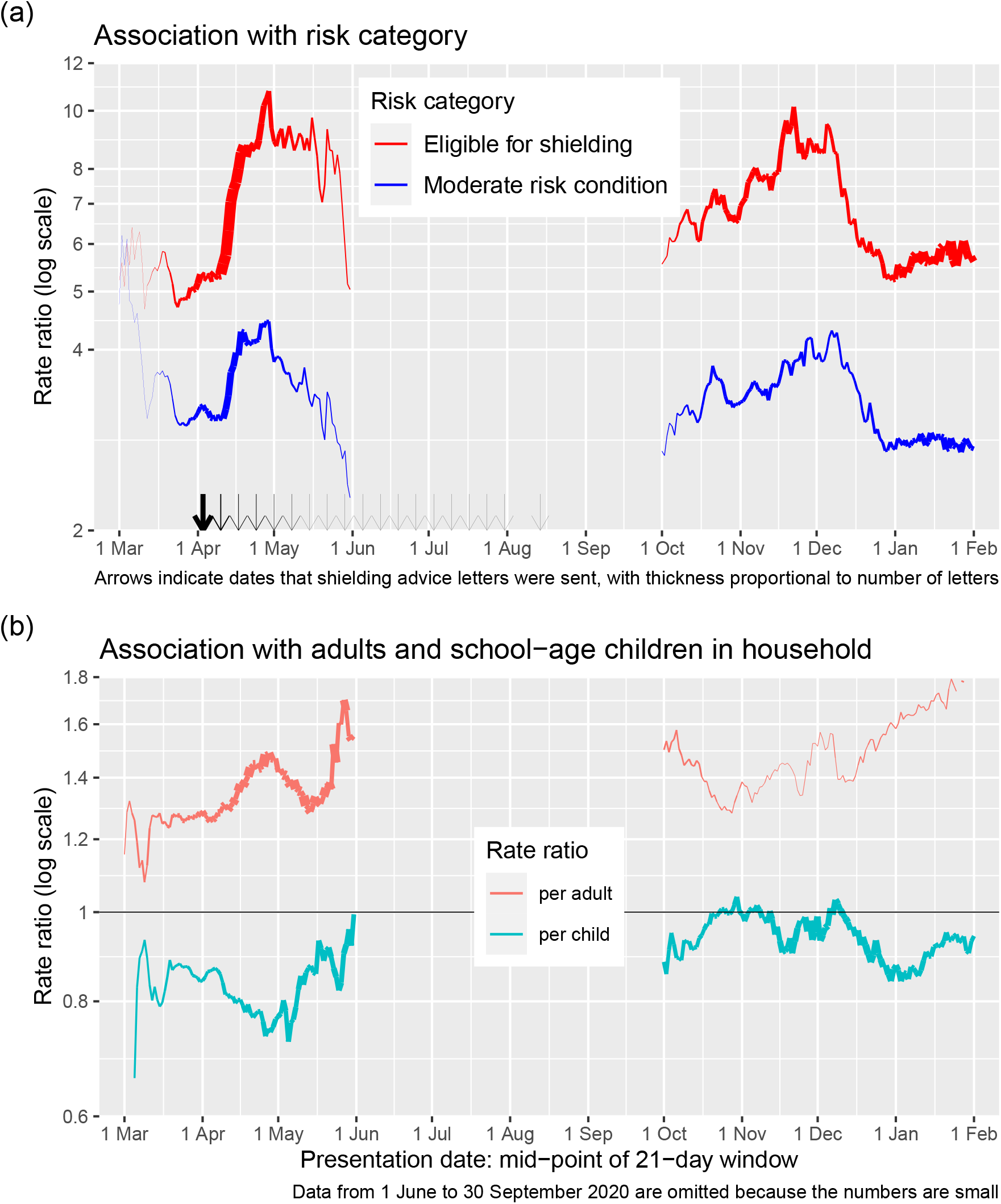
(a) Rate ratios for severe COVID-19 in those not resident in care homes, by risk group and sliding window of presentation dates. (b) Rate ratio per adult and per child in household, in models with number of adults, number of children and SIMD deprivation score as covariates. Line thickness is proportional to precision of estimate.

### Supplementary Tables

**Table S1.** Shielding eligibility cohort by eligibility category and age

|  | Age group |  |  |  | All |
| --- | --- | --- | --- | --- | --- |
|  | 0-39 | 40-59 | 60-74 | 75 or more |  |
| Shielding eligibility category |  |  |  |  |  |
| Solid organ transplant | 1170 (17%) | 2644 (38%) | 2520 (37%) | 537 (8%) | 6872 |
| Specific cancers | 1417 (5%) | 6609 (25%) | 11303 (42%) | 7367 (28%) | 26699 |
| Severe respiratory | 3646 (4%) | 18465 (21%) | 37221 (43%) | 27261 (31%) | 86599 |
| Rare diseases | 2527 (23%) | 3092 (28%) | 2557 (23%) | 2894 (26%) | 11072 |
| On immunosuppressants | 6465 (20%) | 10754 (34%) | 9577 (30%) | 5010 (16%) | 31807 |
| Additional conditions | 6087 (12%) | 13870 (28%) | 15791 (32%) | 13842 (28%) | 49595 |
| All shielding categories | 21312 (10%) | 55434 (26%) | 78969 (37%) | 56911 (27%) | 212644 |

Percentages are row percentages

**Table S2.**
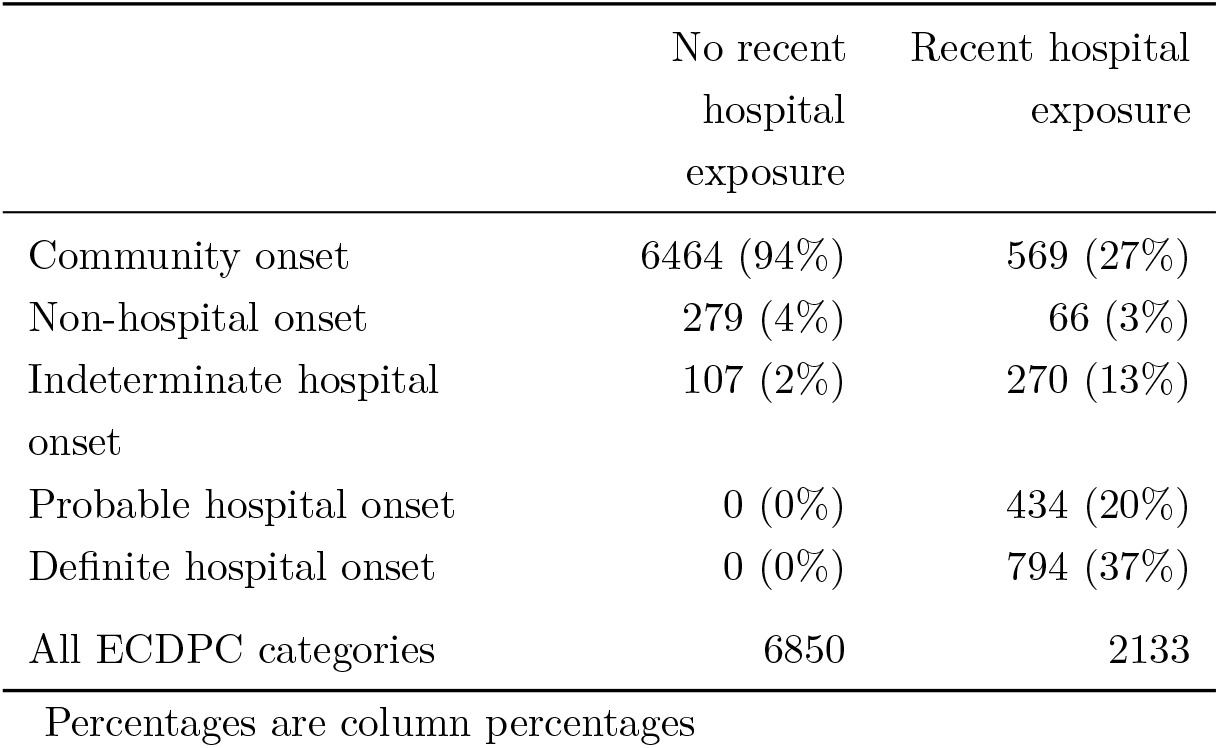
Severe test-positive cases classified by hospital onset status (as defined by ECDPC) and recent inpatient exposure

**Table S3.** Dates of sending advice letters to those eligible for shielding

|  | 3 Apr | 10 Apr | 17 Apr | 24 Apr | 1 May or later | All |
| --- | --- | --- | --- | --- | --- | --- |
| Solid organ transplant | 6674 (97%) | 10 (0%) | 8 (0%) | 22 (0%) | 143 (2%) | 6857 |
| Specific cancers | 17485 (78%) | 241 (1%) | 556 (2%) | 897 (4%) | 3132 (14%) | 22311 |
| Severe respiratory | 77389 (90%) | 424 (0%) | 705 (1%) | 2170 (3%) | 4837 (6%) | 85525 |
| Rare diseases | 916 (11%) | 6363 (76%) | 159 (2%) | 474 (6%) | 444 (5%) | 8356 |
| On immunosuppressants | 13302 (43%) | 9806 (32%) | 4007 (13%) | 1658 (5%) | 2067 (7%) | 30840 |
| Pregnant with heart disease | 18 (41%) | 3 (7%) | 1 (2%) | 8 (18%) | 14 (32%) | 44 |
| Additional conditions | 2287 (5%) | 7290 (17%) | 6626 (16%) | 7554 (18%) | 18024 (43%) | 41781 |
| All shielding categories | 118071 (60%) | 24137 (12%) | 12062 (6%) | 12783 (7%) | 28661 (15%) | 195714 |

Percentages are row percentages

